# A Phase 0 Window of Opportunity Trial of LB100, a Protein Phosphatase 2A Inhibitor, in Patients with Recurrent Gliomas

**DOI:** 10.1101/2025.08.18.25333496

**Authors:** Eric C. Burton, Erin Walker, Lisa Boris, Jennifer Reyes, Kelly Fernandez, Kathleen Wall, Jing Wu, Keith T. Schmidt, William D. Figg, Desmond A. Brown

## Abstract

**Background:** LB100 is a protein phosphatase 2A (PP2A) inhibitor. Glioma models show inhibition of PP2A by LB100 causes cell death. Whether LB100 crosses the human blood brain barrier (BBB) is unknown. We sought to determine the pharmacokinetic (PK) properties of LB100 in human subject gliomas.

**Methods:** A two-stage, phase 0 trial was done. Eligibility required a recurrent adult diffuse type of glioma deemed surgically resectable. In the first stage, 5 patients were pre-surgically dosed with LB100. Resected tumor then underwent PK analysis by LC-MS/MS. If one of five tumors demonstrated a PK response three additional subjects would be enrolled. Pharmacokinetic effect would be declared significant if at least 2 of 8 patients demonstrated a PK response and pharmacodynamic studies would then be performed.

**Results:** Five patients were evaluable. Glioblastoma, (n=2), Astrocytoma IDH-mutant grade 3-4 (n=2), and Oligodendroglioma IDH-mutant, grade 2 (n=1). Mean C_max_ was 146 ng/mL (range: 95-179 ng/mL). Mean plasma half-life (T_1/2_) was 1.2 hours (range: 1.09-1.46 hours). Mean plasma drug exposure (AUC_INF_) was 414 hr*ng/mL (range: 325 to 468 ng*hr/mL. Average concentration of LB100 in tumor was 0.19 nM (range: 0 to 0.67 nM). Average plasma concentration of LB100 was 77.26 nM (range: 30.81 to 132.26 nM). The percent of drug penetration into the brain was 0.31% (range: 0% to 1.04%). The IC_50_ of PP2A is 0.2-0.4 uM; showing drug penetration was inadequate.

**Conclusion:** In this first PK analysis of LB100 in human gliomas there was poor penetration of LB100 into glial tumors.

**Statement of Translational Relevance:** This phase 0 study represents the first pharmacokinetic (PK) analysis of LB100 in human brain cancers. It demonstrates there is poor penetration of LB100 crossing the blood brain barrier (BBB) into central nervous system (CNS) tumors. The ratio of the LB100 concentration in tumor to plasma tissue was much less than one percent, which resulted in a median LB100 tumor tissue concentration of 0.19 nM or 1000-fold less than a previously established minimum IC50 of 0.2uM, targeting the enzyme protein phosphatase 2A. This study highlights the importance of phase 0 studies as a component of new drug development for patients with CNS tumors. The poor brain tumor penetration demonstrated in this investigation will inform decision making regarding future LB100 clinical trials. Unless efforts are successful to reengineer LB100 to increase BBB permeability, upcoming trials will focus on non-CNS tumors.

## INTRODUCTION

Primary gliomas are tumors arising in the central nervous system (CNS) and are largely incurable despite aggressive multimodality therapy consisting of surgical resection, irradiation, and chemotherapy(1). Given the incurability, it should not be surprising that chemotherapeutic options for patients with recurrent glioma are limited, and there is a need to identify effective agents against these tumors.

A problem unique in drug development for tumors located in the central nervous system is the blood-brain barrier (BBB)(2, 3). The BBB is a highly selective semipermeable border of endothelial cells that regulates the transfer of solutes and chemicals between the circulatory system and the CNS, protecting the brain from harmful or unwanted substances in the blood(2). The BBB prevents brain uptake of most pharmaceuticals(4–6). Therefore, drugs being considered for clinical use against CNS tumors must cross the BBB and achieve adequate intratumoral concentration at levels that would affect intracellular mechanisms when given systemically in human subjects.

LB100 is a novel, water-soluble, small molecule inhibitor of protein phosphatase 2A (PP2A) that is an effective growth inhibitor of a variety of CNS and systemic neoplasms in both *in vitro* and *in vivo* models(7, 8). Preclinical studies indicate LB100 has activity as a single agent and is synergistic with other important cytotoxic agents, including temozolomide and ionizing radiation–both standard therapies for many gliomas(7, 9). A completed phase I study of LB100 has established its safety in humans(10). Although it is a polar compound, rodent studies suggest LB100 may have activity in the brain(11). However, whether LB100 crosses the human BBB and at what concentration relative to the plasma level is not known. This study was done to determine the pharmacokinetic (PK) properties of LB100 in human subject glioma tissues.

## PATIENTS AND METHODS

### Patient Selection

All patients were enrolled at the Center for Cancer Research of the National Cancer Institute (NCI), Bethesda, Maryland, USA. Eligible patients included those ≥ 18 years old with histologically confirmed recurrent gliomas that were clinically indicated for resection. Specific eligible tumor types (using WHO criteria) included adult-type diffuse gliomas: Glioblastoma, IDH-wildtype: Astrocytoma, IDH-mutant grade 2-4; Oligodendroglioma, IDH-mutant, and 1p/19q-codeleted grade 2-3. All patients were male and were randomized to a single arm.

Patients had to have a Karnofsky performance score (KPS) of ≥ 60 and adequate organ function including a white blood cell count ≥ 3,000 cells/mm, absolute neutrophil count ≥ 1,500 cells/mm, platelets ≥ 100,000 cells/mm, hemoglobin ≥ 10.0 g/dl, bilirubin ≤ 2 x upper limit of normal (ULN), AST/ALT ≤ 3x ULN and creatinine ≤ 1.7 mg/dl or GFR ≥ 30 ml/min. Patients had to have the ability to understand and the willingness to sign a written informed consent. Exclusion criteria included patients with a history of allergic reaction to LB100 or if taking any other investigational agent, uncontrolled intercurrent illness or prior chemotherapy less than four weeks before study enrollment, patients that were HIV positive on anti-viral therapy, or those on strong CYP450 inhibitors were also ineligible.

The initial screening visit was done within 28 days prior to the scheduled surgery. Basic demographic information and a detailed medical history were collected. Performance status assessment, physical examination, basic laboratory testing as well as baseline MRI were assessed during that visit. Once patients were confirmed eligible, they were enrolled in the study. All enrolled patients finished the study i.e. there was no attrition. The clinical trial was registered on ClinicalTrials.gov (NCT03027388). The study protocol was reviewed and approved by the NCI Institutional Review Board. Written informed consent was obtained from all study participants.

### Study Design and Drug Administration

This was a two-stage, phase 0, open-label, single institution, non-randomized window of opportunity trial. The primary endpoint was to determine the pharmacokinetic profile of LB100 in the setting of recurrent WHO grade 2-4 gliomas.

The recommended phase II starting dose established from a phase I study was 2.33 mg/m2 per day for 3 consecutive days every 3 weeks(10). In this current trial, a single dose of LB100 was infused over two hours (+/− 15 minutes) starting two to four hours before surgery. Because the tumor resection phase of the craniotomy for recurrent gliomas typically lasts 1 to 5 hours, tumor tissue was sampled at multiple time points to allow correlation of tissue drug level with time following infusion.

Blood and plasma samples for PK measurements were also performed on all participants. Samples were collected at the following time points: 1) Pre-dose; 2) End of infusion (2 hours post-start); 3) 30 minutes post LB100 infusion completion; 4) 1 hour post LB-100 infusion completion; 5) 2 hours post LB100 infusion completion; 6) 4 hours post LB100 infusion completion; 7) 8 hours post LB100 infusion completion.

Glioma tissue sampling to detect and quantify LB100 was performed post-dose in each patient, with resection time points from the end of the LB100 infusion recorded. After ensuring adequate diagnostic tissue, remaining tissue was used for PK analysis. Briefly, removed tissues were rinsed with saline immediately (to remove LB100-containing blood and avoid falsely elevated LB100 measurements), then divided, flash-frozen, and stored at −80°C for future batch analysis.

### Statistical Analysis

Up to 25 patients could be enrolled to obtain eight evaluable patients. The intention was to treat an initial five eligible patients, and if one of the five demonstrated a PK response (defined as a binary variable indicating the presence/absence of LB100 in tumor tissues), three additional subjects would be enrolled. Pharmacokinetic effect would be declared to be significant if at least two of the eight patients demonstrated a PK response. Participants whose pathology showed treatment effect or no active tumor would be replaced. Because this was a pilot study, a formal power calculation was not required.

### Bioanalysis from Plasma and Tumor Tissue

LB100 concentration in clinical PK samples (plasma and tumor tissue), was quantified using a validated and robust LC-MS/MS assay developed at the NCI that used LB-105 as an internal standard. This analysis was performed blinded to tumor type. Plasma samples were pre-treated with 50 mM NaOH and tissue samples were homogenized in acetonitrile at a concentration of 100 mg/mL Briefly, samples were mixed with aqueous 10 mM triethylamine (TEA) containing internal standard followed by protein precipitation with 0.5% TEA in acetonitrile. Samples were processed alongside calibration and quality control (QC) standards, ranging from 2-1000 ng/mL (∼7.5-3700 nM) and 3-10,000 pg/mg (∼1.2-3700 nM) for plasma and brain tissue, respectively. Brain tissue concentrations were detectable at values ∼10-fold lower than the lower limit of quantitation (LLOQ; ∼1.2 nM); approximate values for detectable concentrations below LLOQ were extrapolated from the assay calibration curve. The analyte and internal standard were chromatographically separated using a Waters Acquity UPLC BEH C18 (1.7-μm, 2.1x50mm) on a Waters Acuity UPLC system (Waters Corp., Milford, MA). A gradient mobile phase consisting of 5 mM ammonium bicarbonate (aq) and acetronitrile was flowed at a rate of 0.5 to 1 mL/min; the injection volume was 5 uL. After chromatographic separation, the compounds were eluted and detected using a AbSciex QTrap 5500 tandem mass spectrometer in positive ion electrospray mode; MRM transitions were m/z 269.1 ➔ 101.0 and 283.2 ➔ 115.1 for LB-100 and LB-105, respectively.

Plasma PK was analyzed via a non-compartmental approach using Phoenix WinNonlin v8.3 (Certara Corp, Cary, NC) that was validated per FDA 21CFR Part 11 Regulations. The maximum plasma concentration (C_max_) and the time of C_max_ (T_max_) were reported as observed values. A rate constant of the terminal phase, λ_z_, was calculated from the concentration-time curve using uniformly weighted least-squares as the estimation procedure and acceptance criteria of i) r^2^ > 0, ii) includes >3-time points. The area under the concentration-time curve from time 0 to infinity (AUC_INF_) was calculated using the linear-up/log-down trapezoidal method (model type Plasma [200-202]) with extrapolation beyond the last measurable drug concentration (C_last_) calculated via dividing C_last_ by λ_z_ (AUC_INF_ determined to be evaluable if extrapolated AUC beyond the last time point was <25%). Estimated pharmacokinetic parameters included half-life (t_1/2_; inverse of λ_z_), volume of distribution (V_d_; dose divided by product of λ_z_ and AUC_INF_), and clearance (CL; dose divided by AUC_INF_). Drug concentrations in brain tissue samples were reported and % brain penetrance was approximated via the ratio of brain tissue drug concentration to plasma drug concentrations from matched timepoints post-tissue collection.

### Safety and Assessments

After LB100 infusion and craniotomy all patients were evaluated in person 1-3 days following surgery and followed for at least 30 days after infusion of LB100 for toxicities and adverse events (AEs). All adverse events, including clinically significant abnormal findings on laboratory evaluations, regardless of severity, were followed until return to baseline or stabilization of event. Adverse events were documented from the first study intervention (study day 1) through the final safety visit (study day 30). Beyond 30 days only adverse events which were serious and related to the study intervention were recorded. The descriptions and grading scales found in the revised NCI Common Terminology Criteria for Adverse Events (CTCAE) version 5.0 were utilized for AE reporting. An abnormal (grade 2 or greater) laboratory value was recorded in the database as an AE only if the laboratory abnormality was characterized by any of the following: it was thought to be related to study drug, it resulted in discontinuation from the study, it was associated with clinical signs or symptoms, it required treatment or any other therapeutic intervention, it was associated with death or another serious adverse event, including hospitalization, or it was judged by the investigator to be of significant clinical impact. If any abnormal laboratory result was considered clinically significant, the investigator provided details about the action taken with respect to the test drug and the patient’s outcome.

During the first post-operative week after infusion with LB100, an EKG, renal function tests and liver function tests were performed. Two to three weeks following discharge from the hospital, all patients were given a physical exam and additional blood testing. At approximately 30 days post-craniotomy patients were questioned to determine whether any new AEs had occurred, and any ongoing AEs were followed up on. Prior to removal from the study, at approximately 30 days following drug infusion, all subjects completed a final safety evaluation. Data was stored according to HHS, FDA, and NIH regulations.

### Data Availability

Raw data for this study were generated at the NCI. Derived data supporting the findings of this study are available from the corresponding author upon request.

## RESULTS

### Patient Population

The patient demographics and clinical characteristics are summarized in Table 1a,b. For the initial stage of the study, a total of seven patients were enrolled to accrue five evaluable patients. Two patients were excluded due to the surgically resected tissue showing only treatment related changes with no viable tumor tissue in the surgical sample. The tumor types included in this analysis were: glioblastoma, IDH wild-type (n=2), astrocytoma IDH-mutant grade 4 (n=1), astrocytoma IDH-mutant grade 3 (n=1) and oligodendroglioma IDH-mutant, 1p/19q co-deleted grade 2 (n=1). The median patient age was 53 years (range 40-65). All patients were male and completed radiation and chemotherapy prior to enrollment. Three of the five patients received only one treatment regimen before study enrollment (standard radiation and chemotherapy for newly diagnosed tumor). The median KPS was 90 at the time of surgery (range 80-100). The median time from original tumor diagnosis until study enrollment was 51 months (range: 37-181mos). All surgical resections occurred within the designated time interval following the presurgical dose of LB100 (mean 5.68 hours range:5.13 to 6.67 hours).

**Table 1a.** Patient Demographics of all patient participants in the LB100 clinical trial. *Age=censored to protect patient identity. M=male, W=white, KPS=karnofsky performance score, AA=anaplastic astrocytoma, GBM=glioblastoma, G2=grade 2, oligo=oligodendroglioma, IDH-m=IDH mutated, RT/TMZ=standard 6 weeks radiation + temozolomide, bev/nivo= bevacizumab/nivolumab trial

| Patient | Age range at time of enrollment* | Sex | Race | KPS | Tumor diagnosis at time of enrollment | Tumor diagnosis after surgery | Time from original diagnosis to trial entry | Previous treatments | Number of prior regimens |
| --- | --- | --- | --- | --- | --- | --- | --- | --- | --- |
| 1. | XX | M | W | 90 | AA IDH-M | Grade 4, IDH-M astrocytoma | 4yrs 3mo (51months) | RT/TMZ | 1 |
| 2. | XX | M | W | 80 | GBM | GBM | 3yr 10mo (46 months) | RT/TMZ | 1 |
| 3. | XX | M | W | 90 | AA IDH-M | AA IDH-M | 11yr 5mo (137 | RT/TMZ | 1 |
|  |  |  |  |  |  |  | months) |  |  |
| 4. | XX | M | W | 90 | G2 oligo | G2 oligo | 15yr 1mo<br>(181 months) | TMZ, RT | 2 |
| 5. | XX | M | W | 100 | GBM | GBM | 3yr 1mo<br>(37 months) | RT/TMZ,<br>TMZ +/-<br>Veliparib,<br>bev/nivo | 2 |

**Table 1b.**
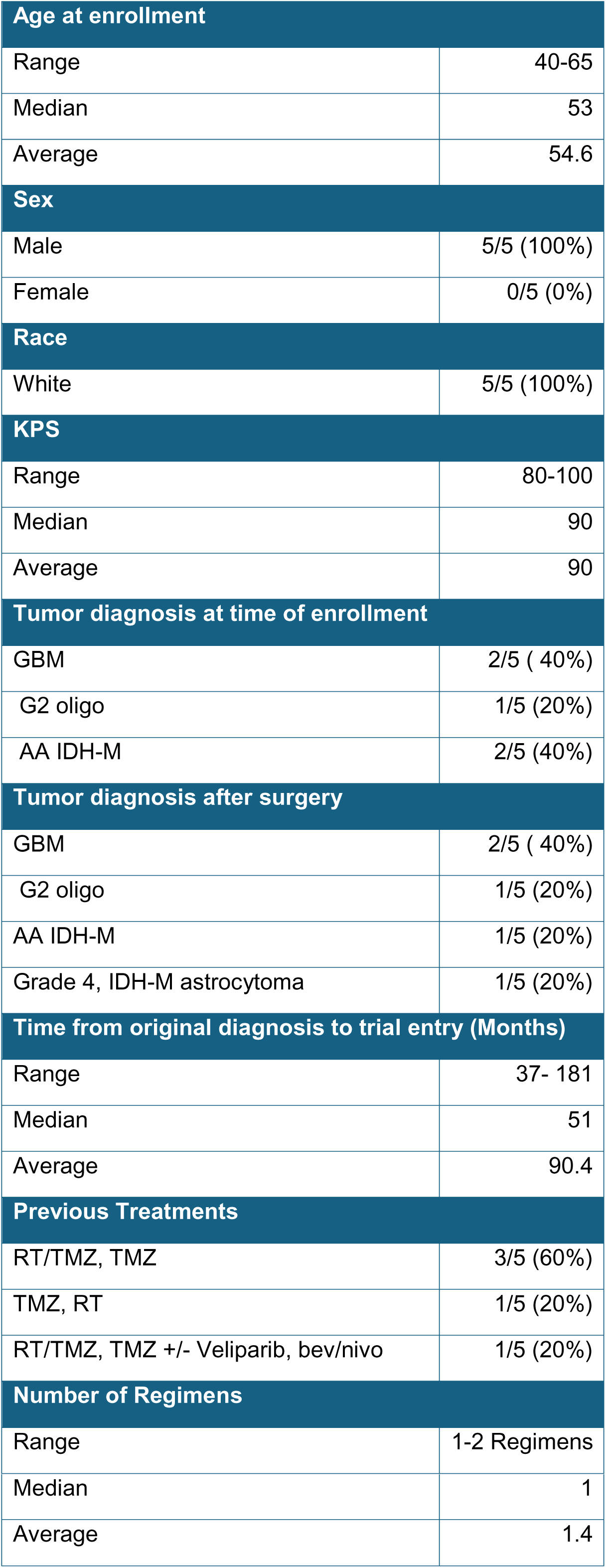
Descriptive statistics of LB100 patient data.

| Age at enrollment |  |
| --- | --- |
| Range | 40-65 |
| Median | 53 |
| Average | 54.6 |
| Sex |  |
| Male | 5/5 (100%) |
| Female | 0/5 (0%) |
| Race |  |
| White | 5/5 (100%) |
| KPS |  |
| Range | 80-100 |
| Median | 90 |
| Average | 90 |
| Tumor diagnosis at time of enrollment |  |
| GBM | 2/5 ( 40%) |
| G2 oligo | 1/5 (20%) |
| AA IDH-M | 2/5 (40%) |
| Tumor diagnosis after surgery |  |
| GBM | 2/5 ( 40%) |
| G2 oligo | 1/5 (20%) |
| AA IDH-M | 1/5 (20%) |
| Grade 4, IDH-M astrocytoma | 1/5 (20%) |
| Time from original diagnosis to trial entry (Months) |  |
| Range | 37- 181 |
| Median | 51 |
| Average | 90.4 |
| Previous Treatments |  |
| RT/TMZ, TMZ | 3/5 (60%) |
| TMZ, RT | 1/5 (20%) |
| RT/TMZ, TMZ +/- Veliparib, bev/nivo | 1/5 (20%) |
| Number of Regimens |  |
| Range | 1-2 Regimens |
| Median | 1 |
| Average | 1.4 |

### Safety

Overall, there were a total of 30 AEs in aggregate from all 7 patients enrolled. Four were rated as serious (grade 3 or 4), with all others being grade 2 or lower. The four AEs rated as serious included one patient with a post-operative headache that resolved with adequate pain management, and one patient who developed anemia and required a transfusion. Two patients had lymphocytopenia. All other AEs were successfully managed or resolved without intervention. No AEs were thought to be related to LB100.

### Plasma Pharmacokinetics

The administered LB100 doses ranged from 4.4 to 5.1 mg (average: 4.74 mg), and the mean time to tumor biopsy after infusion of LB100 was 5.68 hours (5.13 to 6.67 hours). The mean C_max_ was 146 ng/mL (range: 95-179 ng/mL), with the median time to C_max_ after LB100 infusion (T_max_) at 2.10 hours (2.07 to 2.63). The mean plasma half-life (T_1/2_) of LB100 was 1.2 hours (range: 1.09-1.46 hours), comparable to half-life values previously reported(10). Mean plasma drug exposure (AUC_INF_) was 414 hr*ng/mL (range: 325 to 468 ng*hr/mL). Pharmacokinetic parameters with % CV are summarized in **Table 2**.

**Table 2:** Summary of Calculated Pharmacokinetic Parameters (Mean; %CV)

| Drug | C<sub>MAX</sub><br>(ng/mL) | ***T<sub>MAX</sub><br>(hr) | AUC<sub>INF</sub><br>(hr*ng/mL) | T<sub>1/2</sub><br>(hr) | CL<br>(L/hr) | Vd<br>(L) | *Brain<br>Conc<br>(nM) | ** %<br>Brain<br>Pen |
| --- | --- | --- | --- | --- | --- | --- | --- | --- |
| LB100 | 146<br>(22%) | 2.10 (2.07-2.63) | 414 (14%) | 1.27<br>(13%) | 11.7<br>(19%) | 21.7<br>(29%) | 0.19<br>(156%) | 0.31%<br>(159%) |

\*Brain concentrations measured, on average, at 5.7 hr post infusion start.
\*\* Percent brain penetration is the ratio of brain:plasma concentrations (in nM) at the time of biopsy
\*\*\* Median value and range reported instead of mean (% CV)

### Tumor Tissue Pharmacokinetics

The average concentration of LB100 present in the tumor tissue after resection was 0.19 nM or 0.51 pg/mg (range: 0 to 0.67 nM); all LB100 tissue concentration values were below our assay’s LLOQ, and thus were approximated via extrapolation below the assay calibration range. The average plasma concentration of LB100 at the time of biopsy was 77.26 nM or 20.71 ng/mL (range: 30.81 to 132.26 nM). The percent of drug penetration into the brain was 0.31% (range: 0% to 1.04%). The IC_50_ of the LB100 protein target, protein phosphatase 2A (PP2A), in glial cell lines is between 0.2-0.4 uM; therefore, the drug levels in tumor tissue did not achieve a concentration that would be expected to result in a biological response(11–13).

## DISCUSSION

In this phase 0 window of opportunity study, we determined the PK profile of LB100 in human recurrent glioma tumor and plasma tissues. This data represents the first-ever analysis of LB100 in human brain cancer, and it shows there is poor penetration of LB100 crossing the BBB into CNS tumors. The ratio of the LB100 concentration in tumor to plasma tissue was much less than one percent, which resulted in a median LB100 tumor tissue concentration of 0.19 nM or 1000-fold less than a previously established minimum IC50 of 0.2uM, targeting the enzyme protein phosphatase 2A.

These study results evidence the advantages of phase 0 trials. Favorable developmental scenarios for phase 0 studies are in the setting of non-optimized drug formulations, conflicting preclinical data, poor preclinical models, in addition to the prospect of studying pharmacodynamics and pharmacokinetics. All done to inform further drug development. This study was motivated by the uncertainty of LB100 PK properties in humans. There were suggestions that LB100 could penetrate the BBB to a certain degree in rodents. However, the reliability and predictability of animal testing for drug responses and physiological alterations in humans is very low(14). In our model administration of LB100 was shown to significantly decrease PP2A enzymatic activity in the lateral habenula (region of brain with an intact BBB) of mice. Additionally, intracranial glioblastoma mice models were treated with one-time administration of LB100 at an equivalent to maximum tolerated dose in humans. Liver, intracranial tumor, normal brain (animals without tumor implants) was collected and assayed for presence of LB100. Low levels of LB100 were detected in the normal brain and liver from 2 of 3 treated animals and from the implanted tumor in 1 of 3 animals. The mean Cmax in plasma was 1110∼3664 ng/ml. The mean Cmax in the liver and brain were 586∼2548 ng/kg and 17.4∼43.5 ng/kg, respectively (Lixte Biotechnology, Investigators Brochure, unpublished). Given these similarities in drug concentrations (while acknowledging the poor correlation between human and animal models) the findings from this human trial are consistent with pharmacokinetics from animal studies. This makes a false negative result unlikely. Therefore, based on these data, decisions will be made guiding future use of this drug, aligning with the goal of phase 0 studies.

Protein phosphatase 2A is a ubiquitous heterotrimeric serine/threonine phosphatase involved in broad cellular functions such as signal transduction, cell differentiation, regulation of cell cycle, and DNA repair(15). PP2A remains a promising target in glioma(15, 16). Hofstetter et al. demonstrated increased PP2A activity predicts poor outcome in patients with GBM(17). Schramm et al, in a large-scale gene knockdown investigation using diffuse intrinsic pontine glioma cell lines showed the lethal potential of PP2A inhibition (12). Multiple studies have shown PP2A inhibition potentiates the activity of cytotoxic agents including temozolomide and ionizing radiation(13, 18). The mechanism of potentiation appears to be release of cell cycle and mitotic checkpoints allowing dormant cancer cells to continue into mitosis despite acute DNA damage (from cytotoxic agents) resulting in mitotic cell death(13). Regulation of mitosis by PP2A is done by several mechanisms including dephosphorylation and inhibition of polo-like kinase 1 (Plk1) a regulator of DNA damage checkpoint, as well as p53 and AKT(11, 13, 19). In phase 0 studies identifying biomarkers like these is invaluable and critical for assessing drug activity and target modulation. In the event of adequate intratumoral drug levels, biomarkers would have been assessed to ensure that the endpoint of target modulation was in fact observed.

However, while PP2A is an attractive target to sensitize tumor cells to the effect of radiation and chemotherapy, no inhibitor of PP2A is yet FDA approved(20). As such LB100, because of its unique mechanism of action and ability to enhance activity in a broad spectrum of anticancer agents has the potential to be useful for the treatment of many different cancer types beyond CNS tumors (21–23). In some instances, increasing a dose or modifying a drug schedule can be done to improve bioavailability to overcome the limits of the BBB. This is not possible for this LB100 formulation due to toxicity concerns. In the LB100 phase I study 2 of 4 patients had grade 3 increases in creatinine clearance at the dose level above the established safe dose used in this study(10). So LB100 use in CNS tumors will require chemical modification with the aim of introducing BBB permeability. These efforts are currently ongoing.

One potential limitation of pharmacokinetic studies in CNS tumors is sampling bias between contrast and non-contrast enhancing regions of tumor. The contrast-enhancing regions of the tumors often represents the more permeable aspects of the blood-brain tumor barrier. Whereas any drug would be expected to have less penetration into non-contrast-enhancing tumor tissue. We have sought to overcome this limitation by including both contrast and non-contrast regions in our tissue sampling, and in fact we saw no significant difference in drug levels.

This study highlights the importance of phase 0 studies as a component of new drug development for patients with CNS tumors. The poor brain tumor penetration demonstrated in this investigation will inform decision making regarding future LB100 clinical trials. Unless efforts are successful to reengineer LB100 to increase BBB permeability, upcoming trials will focus on non-CNS tumors.

## Data Availability

All data produced in the present work are contained in the manuscript

## Acknowledgments

The authors thank John S Kovach, MD, and Mark Gilbert, MD for providing their valued expertise.

